# Oral Nicotine Pouch Use Among U.S. Middle and High School Students, 2021-2023

**DOI:** 10.64898/2026.01.28.26345040

**Authors:** Haoxiong Sun, Harry Tattan-Birch, Melissa Oldham, Sharon Cox, Sarah E. Jackson

**Author notes:** **Corresponding author:** Haoxiong Sun, BSc, Department of Behavioral Science and Health, University College London, 1-19 Torrington Place London WC1E 7HB, United Kingdom.

## Abstract

**Background:** Oral nicotine pouches (ONPs) have become the second most commonly used nicotine product among U.S. youth. However, little is known about how ONP use is distributed across population subgroups and how strongly it is patterned by use of other tobacco or nicotine products.

**Method:** Data were drawn from the 2021–2023 waves of the National Youth Tobacco Survey (NYTS), a nationally representative survey of U.S. middle and high school students (N = 66,349). We estimated the annual survey-weighted prevalence of current ONP use (≥1 day in the past 30). Using 2023 data (N = 20,174), we estimated prevalence by demographics and other tobacco/nicotine product use, and fitted survey-weighted Poisson regression models to estimate associations of current ONP use with demographics and other nicotine/tobacco product use.

**Results:** In 2023, 1.6% reported current ONP use (95% confidence interval [CI] 1.0–2.1), up from 0.8% in 2021 (95% CI 0.5–1.0). Prevalence in 2023 was higher among males (2.3%, 95% CI 1.5–3.1) than females (0.8%, 95% CI 0.4–1.3), and among students who used any other tobacco or nicotine product (13.4%, 95% CI 10.3–16.5) than those who did not (0.4%, 95% CI 0.1–0.8). In fully adjusted models, use of nicotine products other than cigarettes or e-cigarettes showed the strongest association with ONP use (APR 21.1, 95% CI 13.0–34.0), followed by cigarette smoking (APR 2.0, 95% CI 1.1–3.7) and e-cigarette use (APR 1.9, 95% CI 0.8–4.5). Most current ONP users also used other tobacco/nicotine products (75.0%), though 16.6% reported no lifetime use of other products.

**Conclusions:** Overall ONP prevalence among U.S. youth remains low but is increasing. While ONP use is largely concentrated in youth who use other nicotine/tobacco products, it is also increasing among adolescents who are otherwise nicotine-naïve. These findings highlight the need for continued monitoring and targeted regulations that balance harm reduction for people who smoke against the risk of expanding nicotine dependence among youth.

**Key Points:** 

**Question:** What are the prevalence, demographic patterns, and tobacco/nicotine co-use profiles of oral nicotine pouch users among US middle and high school students?

**Findings:** In this cross-sectional study of 66,349 students from the 2021-2023 National Youth Tobacco Survey, current oral nicotine pouch use doubled from 0.8% in 2021 to 1.6% in 2023. Most users also used other tobacco or nicotine products, but the proportion of current users with no lifetime use of other products increased from 7.4% to 16.6%.

**Meaning:** Although oral nicotine pouch use among US youth remains low, increasing uptake among adolescents without prior tobacco or nicotine exposure suggests a need for targeted prevention efforts alongside continued surveillance.

## Introduction

Oral nicotine pouches (ONPs) are nicotine-containing products placed between the gum and cheek, allowing nicotine absorption through the oral mucosa [1–3]. Unlike other products, ONPs can be used discreetly, including in places where smoking or vaping is prohibited and produce no smoke or aerosol. Similar to vaping products, they are available in multiple flavours and nicotine strengths. Over recent years, there has been rapid market expansion and growing attention from public health and regulators [1, 2, 4–6].

Regulatory approaches to ONPs vary widely across settings. In the United States (U.S.), nicotine pouches are regulated as tobacco products and are subject to FDA premarket review, with marketing authorisation granted for some products [7, 8]. In contrast, in some other jurisdictions (e.g., the United Kingdom), nicotine pouches fall within regulatory gaps, with no age-of-sale restrictions, which has contributed to concerns about youth access and youth-targeted marketing [2, 9].

In the U.S., ONP use among adolescents remains lower than e-cigarette use, but national surveillance indicates an upward trajectory [9]. The annual National Youth Tobacco Survey (NYTS) captures data on use of different nicotine products among middle and high school students. In 2024, nicotine pouches were the second most commonly used tobacco product among U.S. middle and high school students (after e-cigarettes), overtaking cigarettes [9]. However, overall prevalence estimates may obscure important heterogeneity in user profiles. In particular, there is limited evidence distinguishing between ONP users with a history of smoking or other tobacco and nicotine product use and those reporting no prior use of such products.

Subgroup and co-use patterns are important for interpreting the public health implications of ONP use among youth and for targeting prevention efforts. On one hand, ONPs deliver nicotine and may contribute to dependence among adolescents who would otherwise have remained nicotine-free [3, 10, 11]. On the other hand, if ONPs attract youth who would otherwise have initiated smoking, they may offer a relative harm reduction benefit given their substantially lower toxicant profile compared with combustible cigarettes [2, 3, 12, 13]. Emerging evidence suggests ONP uptake may be concentrated within particular subgroups, especially young males, and usually occurs alongside other tobacco or nicotine product use [14, 15]. Although top-line estimates of the prevalence of ONP use within the NYTS are published annually [9], more detailed analysis of the microdata can be undertaken to provide further insights. This study aimed to provide a comprehensive analysis of ONP use among U.S. middle- and high-school students using NYTS data collected between 2021 and 2023. Specifically, we addressed the following research questions:

1. What is the prevalence of current, frequent, and ever ONP use among U.S. middle- and high-school students, overall and by sex, grade, race/ethnicity, and use of other tobacco or nicotine products.
2. What are the demographic and tobacco or nicotine use profiles of those reporting current, former, ever, and never ONP use.
3. What factors are independently associated with current ONP use and demographic characteristics and use of other tobacco or nicotine products.
4. How has ONP prevalence changed from 2021 to 2023 across key subgroups.

## Method

### Pre-registration

Analyses followed a pre-registered protocol (OSF: https://osf.io/x54t2). We report three deviations from the protocol: (1) demographic characteristics were analyzed using school level (middle vs. high school) rather than age; (2) the definition of other nicotine/tobacco products was expanded from 3 to 11 categories (listed below); and (3) we added a post-hoc descriptive analysis to characterise the tobacco or nicotine product use histories of current ONP users.

### Design and Sample

Data were drawn from the NYTS, an annual, nationally representative survey of U.S. middle- and high-school students (grades 6–12) conducted by the CDC and FDA [16]. The NYTS employs a stratified, three-stage cluster sampling design. Questionnaires were administered to students during a regular class period [17].

The most recent NYTS was conducted in 2024. However, while top-line estimates have been published [9], the microdata are not currently available and it is not clear when (or whether) they will be. This study therefore used data from 2021 (N = 19142), 2022 (N = 27033), and 2023 (N = 20174) waves. Analyses of prevalence, user profiles, and associations (Tables 1–3) were based on the 2023 wave (N = 20,174) to provide the most up-to-date estimates. Trend analyses used data from all three waves (2021–2023). Participants who provided information on ONP use were eligible for analysis. NYTS datasets are fully de-identified and publicly available; therefore, this secondary analysis did not require institutional ethical review.

**Table 1.** Weighted prevalence of oral nicotine pouch (ONP) use among U.S. middle and high school students, National Youth Tobacco Survey 2023.

|  |  | Prevalence % (95% CI) |  |  |
| --- | --- | --- | --- | --- |
| Characteristic | Unweighted<br>N | Ever ONP use<sup>a</sup> | Current ONP use<sup>b</sup> | Current frequent ONP use<sup>c</sup> |
| Overall |  |  |  |  |
| All students | 20174 | 2.7% (2.1-3.4) | 1.6% (1.0-2.1) | 0.4% (0.3-0.6) |
| Demographic characteristics |  |  |  |  |
| Grade |  |  |  |  |
| Middle school | 10192 | 1.7% (0.7-2.7) | 1.1% (0.3-2.0) | 0.4% (0.1-0.6) |
| High school | 9912 | 3.5% (2.7-4.2) | 1.8% (1.1-2.5) | 0.5% (0.2-0.8) |
| Sex |  |  |  |  |
| Female | 10007 | 1.8% (1.3-2.4) | 0.8% (0.4-1.3) | 0.1% (0.0-0.2) |
| Male | 10053 | 3.6% (2.6-4.6) | 2.3% (1.5-3.1) | 0.8% (0.5-1.2) |
| Race/ethnicity |  |  |  |  |
| Hispanic | 5539 | 2.9% (1.7-4.1) | 2.0% (0.9-3.2) | 0.5% (0.1-0.9) |
| Non-Hispanic Other | 2846 | 0.8% (0.3-1.3) | 0.5% (0.1-0.8) | 0.2% (0.0-0.3) |
| Non-Hispanic Black | 2474 | 1.4% (0.5-2.2) | 1.1% (0.4-1.8) | 0.3% (0-0.7) |
| Non-Hispanic White | 8821 | 3.2% (2.4-4.0) | 1.5% (0.8-2.2) | 0.5% (0.2-0.7) |
| Tobacco or nicotine use |  |  |  |  |
| E-cigarette use |  |  |  |  |
| Current | 1280 | 20.0% (15.0-25.1) | 11.5% (7.7-15.3) | 3.8% (1.9-5.7) |
| Not current | 18878 | 1.5% (0.9-2.0) | 0.8% (0.3-1.3) | 0.2% (0.1-0.3) |
| Cigarette use |  |  |  |  |
| Current | 385 | 39.7% (31.3-48.1) | 29.7% (22.2-37.3) | 10.4% (5.5-15.3) |
| Not current | 19784 | 2.0% (1.5-2.5) | 1.0% (0.6-1.5) | 0.3% (0.1-0.4) |
| Other tobacco or nicotine product use |  |  |  |  |
| Current | 748 | 35.7% (28.2-43.1) | 28.1% (22.0-34.2) | 9.6% (5.9-13.3) |
| Not current | 19425 | 1.5% (1.1-1.9) | 0.6% (0.2-0.9) | 0.1% (-0.0-0.2) |
| Any tobacco or nicotine product |  |  |  |  |
| Current | 1694 | 21.5% (17.6-25.4) | 13.4% (10.3-16.5) | 4.2% (2.5-5.9) |
| Not current | 18462 | 0.9% (0.6-1.3) | 0.4% (0.1-0.8) | 0.1% (0.0-0.2) |

CI=Confidence interval. ONP=Oral nicotine pouch
a. *Ever ONP use* combines current frequent, current, and former users.
b. *Current use* = use on $\geq 1$ of the past 30 days.
c. *Current frequent use* = use on $\geq 20$ of the past 30 days.
Corresponding stratified data for 2021 and 2022 are provided in Supplementary Tables 1-2.

**Table 2.** Factors associated with current ONP use among U.S. middle and high school students, National Youth Tobacco Survey 2023.

| **Exposure** | **Unadjusted PR (95% CI)<sup>a</sup>**<br>*Crude* | **Demographic-adjusted PR (95% CI)<sup>b</sup>**<br>*Model 1* | **Fully-adjusted PR (95% CI)<sup>c</sup>**<br>*Model 2* |
| --- | --- | --- | --- |
| **Demographics** |  |  |  |
| Gender |  |  |  |
| Male vs Female | 2.67 (1.76–4.06) | 2.67 (1.68–4.22) | 2.65 (1.80–3.89) |
| School level |  |  |  |
| High vs Middle school | 1.61 (0.69–3.73) | 1.71 (0.71–4.15) | 1.23 (0.57–2.65) |
| Race/ethnicity |  |  |  |
| Hispanic vs Non Hispanic-White | 1.24 (0.62–2.47) | 1.10 (0.52–2.35) | 0.95 (0.46–1.95) |
| Non Hispanic-Other vs Non Hispanic-White | 0.29 (0.13–0.63) | 0.28 (0.12–0.61) | 0.36 (0.13–1.03) |
| Non Hispanic-Black vs Non Hispanic-White | 0.65 (0.34–1.25) | 0.66 (0.34–1.31) | 0.69 (0.39–1.21) |
| **Non-ONP Tobacco or nicotine use** |  |  |  |
| E-cigarette use |  |  |  |
| Currently use vs other | 12.89 (6.06–27.42) | 13.86 (6.39–30.04) | 1.91 (0.81–4.50) |
| Cigarette use |  |  |  |
| Currently use vs other | 24.90 (16.12–38.47) | 23.50 (15.78–35.01) | 2.04 (1.12–3.71) |
| Other nicotine/tobacco use<sup>d</sup> |  |  |  |
| Current use vs other | 42.38 (24.64–72.91) | 40.40 (24.45–66.78) | 21.05 (13.03–33.99) |

PR=Prevalence ratio, derived from Poisson regression accounting for complex survey design and weighting.
CI=Confidence interval. ONP=Oral nicotine pouch
*a.*Crude models were unadjusted;
*b.*Model 1 adjusted for demographic factors (sex, school level, and race/ethnicity);
*c.*Model 2 additionally adjusted for current cigarette, e-cigarette, and other nicotine/tobacco use.
*d.*Sensitivity analysis excluding snus from the other nicotine/tobacco use composite variable is shown in Supplementary Table 3.

**Table 3.**
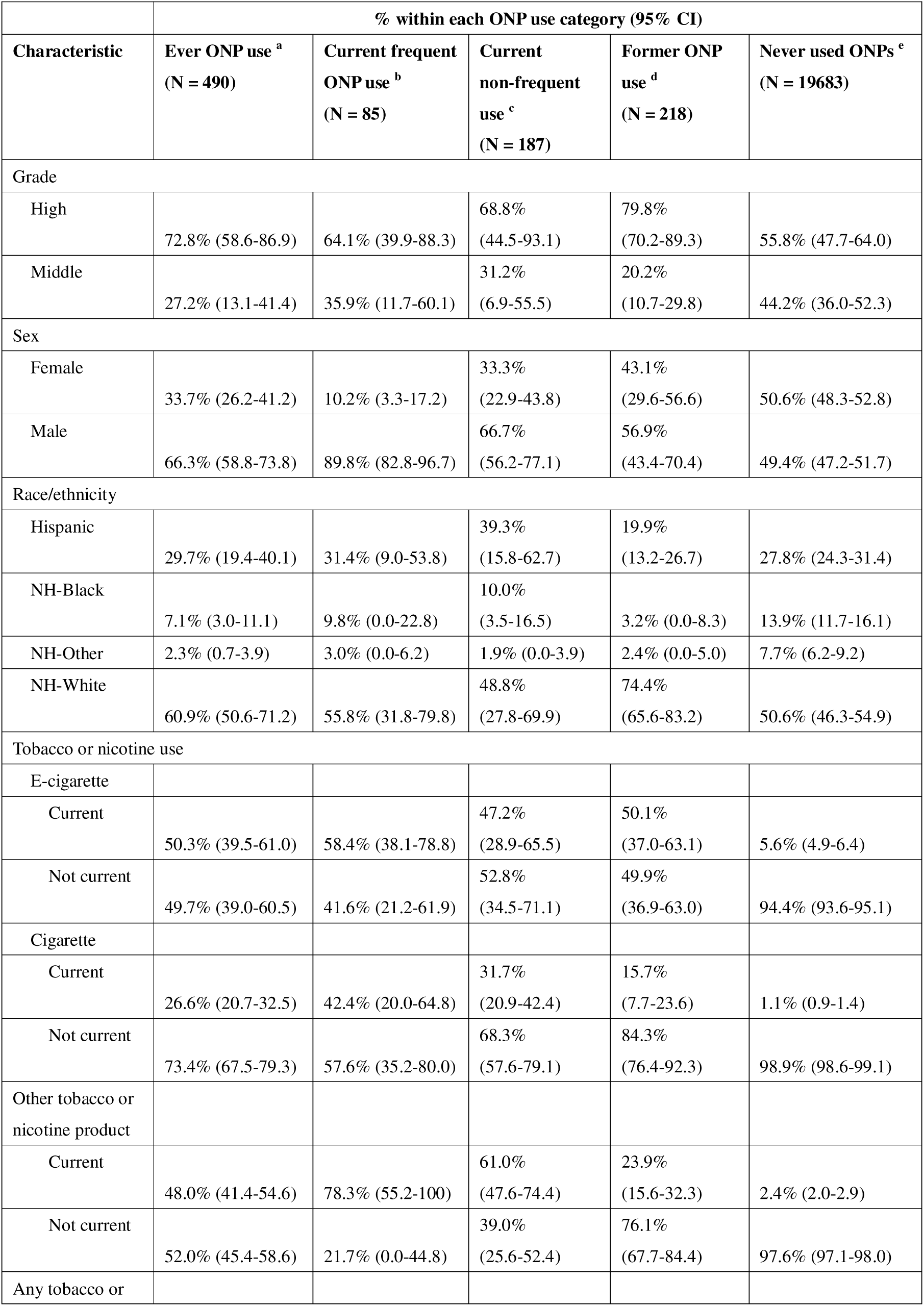
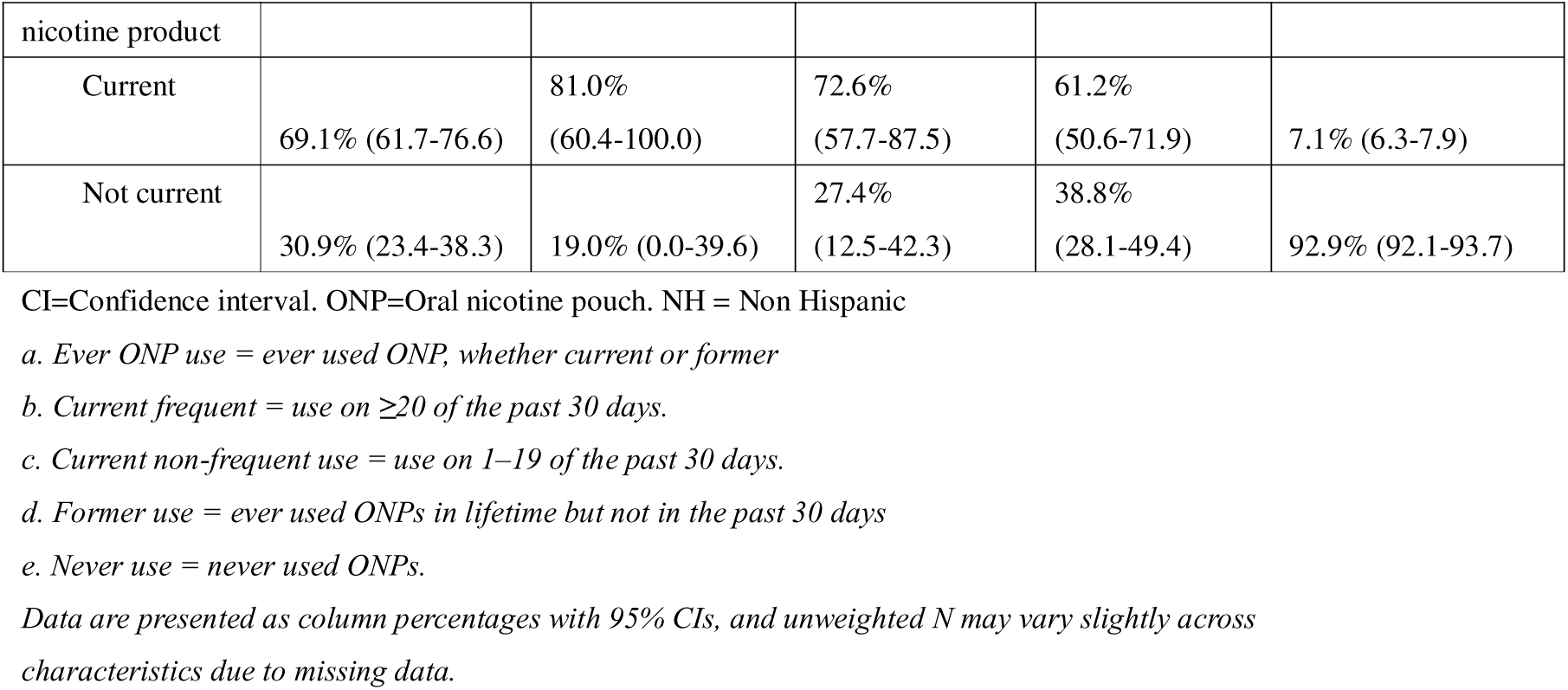
Distribution of demographic and tobacco or nicotine product characteristics across ONP use categories, National Youth Tobacco Survey 2023.

| Characteristic | % within each ONP use category (95% CI) |  |  |  |  |
| --- | --- | --- | --- | --- | --- |
|  | Ever ONP use <sup>a</sup><br>(N = 490) | Current frequent<br>ONP use <sup>b</sup><br>(N = 85) | Current<br>non-frequent<br>use <sup>c</sup><br>(N = 187) | Former ONP<br>use <sup>d</sup><br>(N = 218) | Never used ONPs <sup>e</sup><br>(N = 19683) |
| Grade |  |  |  |  |  |
| High | 72.8% (58.6-86.9) | 64.1% (39.9-88.3) | 68.8%<br>(44.5-93.1) | 79.8%<br>(70.2-89.3) | 55.8% (47.7-64.0) |
| Middle | 27.2% (13.1-41.4) | 35.9% (11.7-60.1) | 31.2%<br>(6.9-55.5) | 20.2%<br>(10.7-29.8) | 44.2% (36.0-52.3) |
| Sex |  |  |  |  |  |
| Female | 33.7% (26.2-41.2) | 10.2% (3.3-17.2) | 33.3%<br>(22.9-43.8) | 43.1%<br>(29.6-56.6) | 50.6% (48.3-52.8) |
| Male | 66.3% (58.8-73.8) | 89.8% (82.8-96.7) | 66.7%<br>(56.2-77.1) | 56.9%<br>(43.4-70.4) | 49.4% (47.2-51.7) |
| Race/ethnicity |  |  |  |  |  |
| Hispanic | 29.7% (19.4-40.1) | 31.4% (9.0-53.8) | 39.3%<br>(15.8-62.7) | 19.9%<br>(13.2-26.7) | 27.8% (24.3-31.4) |
| NH-Black | 7.1% (3.0-11.1) | 9.8% (0.0-22.8) | 10.0%<br>(3.5-16.5) | 3.2% (0.0-8.3) | 13.9% (11.7-16.1) |
| NH-Other | 2.3% (0.7-3.9) | 3.0% (0.0-6.2) | 1.9% (0.0-3.9) | 2.4% (0.0-5.0) | 7.7% (6.2-9.2) |
| NH-White | 60.9% (50.6-71.2) | 55.8% (31.8-79.8) | 48.8%<br>(27.8-69.9) | 74.4%<br>(65.6-83.2) | 50.6% (46.3-54.9) |
| Tobacco or nicotine use |  |  |  |  |  |
| E-cigarette |  |  |  |  |  |
| Current | 50.3% (39.5-61.0) | 58.4% (38.1-78.8) | 47.2%<br>(28.9-65.5) | 50.1%<br>(37.0-63.1) | 5.6% (4.9-6.4) |
| Not current | 49.7% (39.0-60.5) | 41.6% (21.2-61.9) | 52.8%<br>(34.5-71.1) | 49.9%<br>(36.9-63.0) | 94.4% (93.6-95.1) |
| Cigarette |  |  |  |  |  |
| Current | 26.6% (20.7-32.5) | 42.4% (20.0-64.8) | 31.7%<br>(20.9-42.4) | 15.7%<br>(7.7-23.6) | 1.1% (0.9-1.4) |
| Not current | 73.4% (67.5-79.3) | 57.6% (35.2-80.0) | 68.3%<br>(57.6-79.1) | 84.3%<br>(76.4-92.3) | 98.9% (98.6-99.1) |
| Other tobacco or<br>nicotine product |  |  |  |  |  |
| Current | 48.0% (41.4-54.6) | 78.3% (55.2-100) | 61.0%<br>(47.6-74.4) | 23.9%<br>(15.6-32.3) | 2.4% (2.0-2.9) |
| Not current | 52.0% (45.4-58.6) | 21.7% (0.0-44.8) | 39.0%<br>(25.6-52.4) | 76.1%<br>(67.7-84.4) | 97.6% (97.1-98.0) |
| Any tobacco or |  |  |  |  |  |

| nicotine product |  |  |  |  |  |
| --- | --- | --- | --- | --- | --- |
| Current | 69.1% (61.7-76.6) | 81.0%<br>(60.4-100.0) | 72.6%<br>(57.7-87.5) | 61.2%<br>(50.6-71.9) | 7.1% (6.3-7.9) |
| Not current | 30.9% (23.4-38.3) | 19.0% (0.0-39.6) | 27.4%<br>(12.5-42.3) | 38.8%<br>(28.1-49.4) | 92.9% (92.1-93.7) |

CI=Confidence interval. ONP=Oral nicotine pouch. NH = Non Hispanic
*a. Ever ONP use = ever used ONP, whether current or former*
*b. Current frequent = use on $\geq 20$ of the past 30 days.*
*c. Current non-frequent use = use on 1–19 of the past 30 days.*
*d. Former use = ever used ONPs in lifetime but not in the past 30 days*
*e. Never use = never used ONPs.*
*Data are presented as column percentages with 95% CIs, and unweighted N may vary slightly across characteristics due to missing data.*

## Measures

### Oral Nicotine Pouch (ONP) Use

ONP use was assessed using two questions: “Have you ever used a nicotine pouch, even just one time?” and “During the past 30 days, on how many days did you use a nicotine pouch?”

For the current study, we defined ONP use in two ways. For prevalence estimation and modeling, we created three non-mutually exclusive categories: ever ONP use (used ONPs at least once), current ONP use (used ONPs on ≥1 day in the past 30 days), and current frequent ONP use (used ONPs on ≥20 days in the past 30 days). For the user profile analysis, we created four mutually exclusive categories: current frequent use (≥20 of the past 30 days), current non-frequent use (1–19 of the past 30 days), former use (ever used but 0 days in the past 30 days), and never use (never used ONPs).

### Demographic Factors

We included three demographic characteristics: sex, school level (middle school vs. high school), and race/ethnicity. Based on responses to Hispanic origin and race questions, participants were categorized into four groups consistent with previous research: Hispanic, Non-Hispanic Black, Non-Hispanic White, and Non-Hispanic Other [22, 26, 27].

### Non-ONP Tobacco or nicotine Product Use

Use of 11 non-ONP products was recorded: (1) e-cigarettes; (2) cigarettes; (3) cigars (cigars, cigarillos, or little cigars); (4) smokeless tobacco (chewing tobacco, snuff, or dip); (5) snus; (6) hookahs; (7) pipe tobacco; (8) bidis; (9) heated tobacco products; (10) dissolvable tobacco; and (11) roll-your-own cigarettes. For each product, we classified status as either current use (≥1 day in the past 30 days), former use (ever use but no use in the past 30 days), or never use.

Cigarette (including roll-your-own cigarettes) and e-cigarette use status were used in the analysis as separate measures. In addition, two composite measures were defined: other non-ONP tobacco or nicotine use (aggregating products 3–10, excluding cigarettes and e-cigarettes) and any non-ONP tobacco or nicotine use (aggregating all 11 products). For both composites, status was classified as current (current use of at least one included product), former (ever use of at least one included product, but no current use of any included product) or never use.

### Statistical Analysis

Analyses were conducted in R (version 4.5.0; R Foundation for Statistical Computing, Vienna, Austria). We excluded participants with missing data on ONP use. Missing data on other variables were excluded on a per-analysis basis. All analyses incorporated survey design features by applying student sampling weights, primary sampling units, and stratification variables using the survey package. All results were reported alongside 95% confidence intervals (CIs).

To address Aim 1, we estimated the prevalence of ever, current, and current frequent ONP use in 2023, overall and within each demographic (sex, school level, race/ethnicity) and non-ONP tobacco or nicotine use status.

For Aim 2, we estimated the percentages of individuals in each demographic and non-ONP tobacco or nicotine use subgroup in 2023 within each mutually exclusive ONP use category.

For Aim 3, we fitted Poisson regression models with a log link and robust variance to estimate prevalence ratios (PRs) for current ONP use (vs. no current use) in 2023, using three specifications: crude (unadjusted) models, Model 1 adjusted for demographic factors (sex, school level, and race/ethnicity), and Model 2 further adjusted for current cigarette, e-cigarette, and other tobacco or nicotine product use. An unplanned sensitivity analysis examined the association between other tobacco or nicotine product use and current ONP use with and without snus (oral tobacco pouches) included in the composite measure, to explore whether this association may have been driven by misreporting of ONP use as snus use.

For Aim 4, we estimated the prevalence of current ONP use in 2021, 2022, and 2023 by demographic and non-ONP nicotine/tobacco use subgroups.

## Results

### Current prevalence and subgroup differences

In 2023, ONP use remained relatively uncommon among U.S. middle and high school students (Table 1). Overall, 2.7% (95% CI: 2.1–3.4) of students reported ever using ONPs, 1.6% (95% CI: 1.0–2.1) reported current use (≥1 day in the past 30 days), and 0.4% (95% CI: 0.3–0.6) reported frequent use (≥20 days).

Prevalence differed across demographic subgroups, notably by sex. Male students (2.3%, 95% CI: 1.5–3.1) had nearly triple the prevalence of current ONP use compared to female students (0.8%, 95% CI: 0.4–1.3). Prevalence also appeared higher among high school students (1.8%, 95% CI: 1.1–2.5) than middle school students (1.1%, 95% CI: 0.3–2.0), though their 95% CIs overlapped. By race and ethnicity, point estimates were highest for Hispanic (2.0%, 95% CI: 0.9–3.2) and non-Hispanic White students (1.5%, 95% CI: 0.8–2.2).

Differences were more pronounced when stratified by other tobacco or nicotine product use. Among current cigarette smokers, 29.7% (95% CI: 22.2–37.3) reported current ONP use, compared with only 1.0% (95% CI: 0.6–1.5) among non-smokers. Current e-cigarette users (11.5%, 95% CI: 7.7–15.3) and users of other tobacco or nicotine products (28.1%, 95% CI: 22.0–34.2) also reported higher ONP use compared with their non-user counterparts (0.8%, 95% CI: 0.3–1.3 and 0.6%, 95% CI: 0.2–0.9, respectively). Similarly, students who currently used any tobacco or nicotine product reported markedly higher ONP use (13.4%, 95% CI: 10.3–16.5) than those who did not (0.4%, 95% CI: 0.1–0.8).

In survey-weighted Poisson models (**Table 2**) adjusting for demographics and non-ONP tobacco or nicotine use, females had lower prevalence of current ONP use than males (fully-adjusted prevalence ratio [APR] = 0.38, 95% CI 0.26–0.56).

Associations with other nicotine product use were strong, with current use of other non-ONP tobacco or nicotine products showing the largest association (APR = 21.05, 95% CI 13.03–33.99), followed by current cigarette smoking (APR = 2.04, 1.12–3.71), and current e-cigarette use (APR = 1.91, 0.81–4.50). When we excluded snus from the composite measure of use of other non-ONP tobacco or nicotine products, the association was attenuated but remained strong (APR = 11.96, 95% CI: 4.94–28.99; **Supplementary Table 3**).

### Trends in ONP use over time (2021–2023)

Between 2021 and 2023, ONP use among U.S. students increased (Figure 1; Supplementary Tables 1, 2). The prevalence of current use rose from 0.8% (95% CI: 0.5–1.0) in 2021 to 1.6% (95% CI: 1.0–2.1) in 2023, and current frequent use increased from 0.1% (95% CI: 0.1–0.2) to 0.4% (95% CI: 0.3–0.6) (Supplementary Tables 1, 2). Increases were observed across most demographic subgroups. Although absolute prevalence of ONP use remains low, these descriptive trends suggest increasing ONP use among U.S. youth over this period.

**Figure 1.**
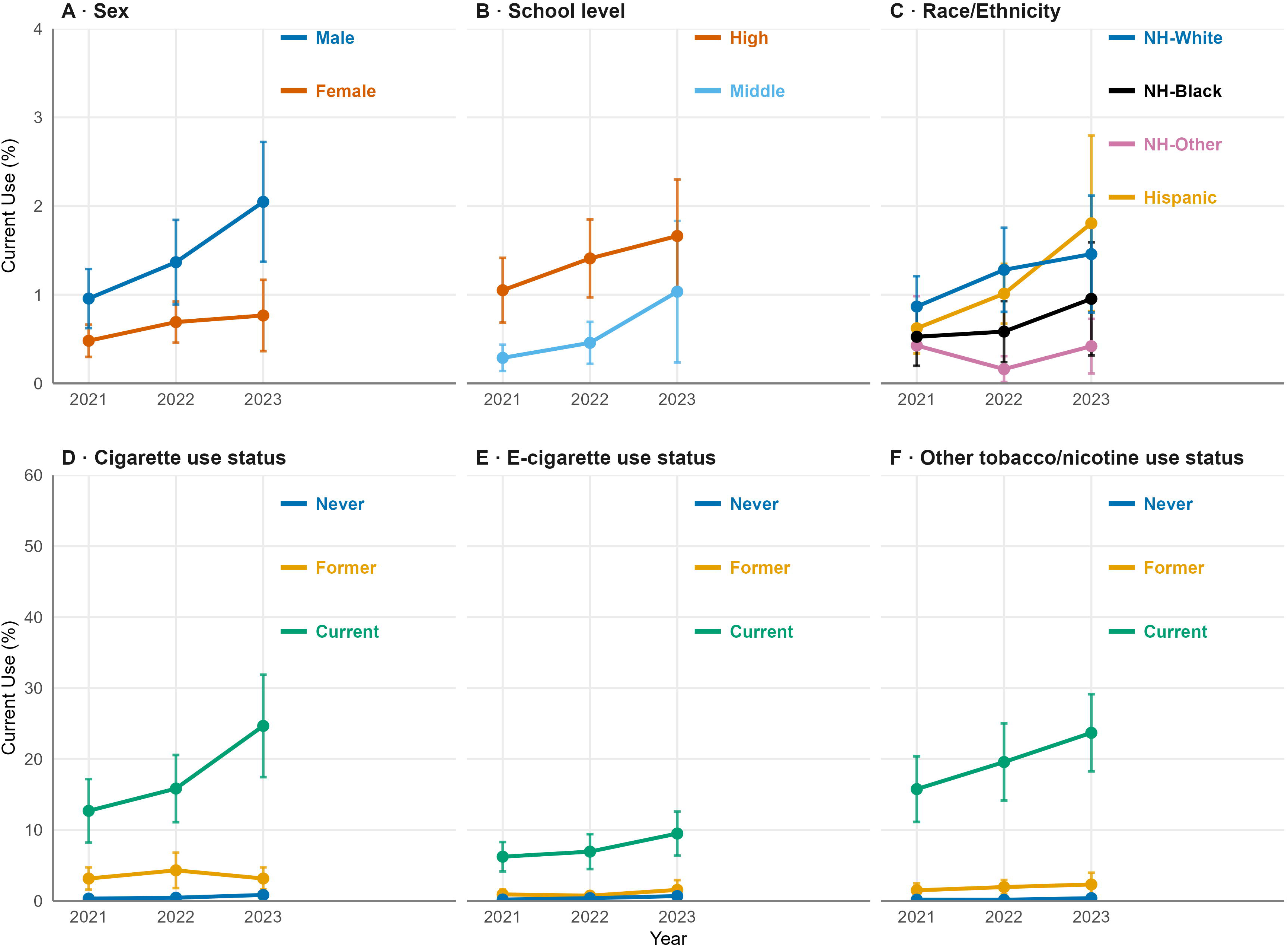
Prevalence of current oral nicotine pouch (ONP) use among U.S. students, National Youth Tobacco Survey 2021–2023. Panels A–C show prevalence by sex, school level, and race/ethnicity; Panels D–F by cigarette, e-cigarette, and other non-ONP tobacco or nicotine product use status. Current = ≥1 day in the past 30 days; former = ever but not current; never = no lifetime use. Estimates are survey-weighted with 95% CIs.

### Profile of current ONP users

**Table 3** summarises the demographic and tobacco or nicotine use profile by ONP use category in 2023. By school level, high school students made up 72.8% of ever ONP users and 64.1% of people who currently use ONPs frequently, compared with 55.8% among those who never used ONPs. Differences by sex were more pronounced: for example, 89.8% of frequent current ONP users were male, versus an approximately even split among those who had never used ONPs (49.4% male; 50.6% female). Patterns by race/ethnicity were harder to interpret in this descriptive comparison because confidence intervals were wide for some ONP use categories.

Most notably, use of other tobacco or nicotine products was common among people who used ONP. For example, 81.0% of frequent current ONP users reported current use of any tobacco or nicotine product, compared with 7.1% among those who had never used ONPs, with intermediate percentages for current non-frequent users (72.6%) and former ONP users (61.2%). Similar contrasts were observed for current e-cigarette use, current cigarette use, and current other tobacco or nicotine product use.

**Figure 2** shows trends in product use status among current ONP users from 2021–2023. Current use of any other tobacco or nicotine product was high in 2021 (85.4%) and 2022 (86.3%) and declined in 2023 (75.0%). Across specific product categories, current e-cigarette use decreased from 64.2% (2021) to 62.7% (2022) and 50.4% (2023), while current cigarette use increased from 30.4% to 31.6% and 34.7%. Current use of other tobacco or nicotine products was 65.4% in 2021, 72.9% in 2022, and 66.0% in 2023. Over the same period, exclusive current ONP use increased, including exclusive users with prior lifetime use of other products (7.2%, 4.0%, and 8.4%) and those with no lifetime use of other products (7.4%, 9.8%, and 16.6%) in 2021, 2022, and 2023, respectively.

**Figure 2.**
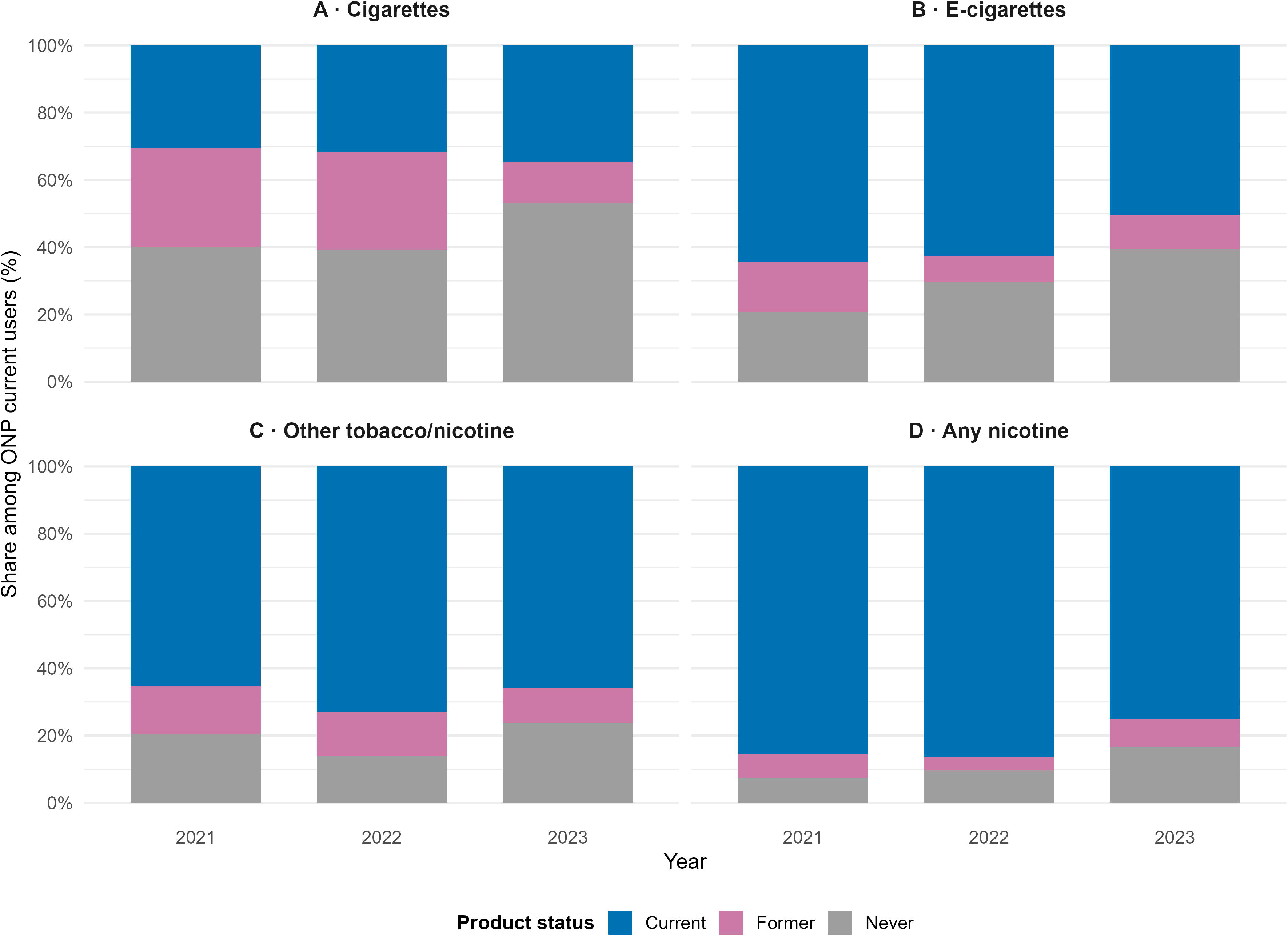
Product use status among current oral nicotine pouch (ONP) users, National Youth Tobacco Survey 2021–2023. Panels A–C show status for cigarettes, e-cigarettes, and other non-ONP tobacco or nicotine products; Panel D shows “any non-ONP tobacco or nicotine” aggregated across products. Bars represent weighted percentages that sum to 100% within year.

## Discussion

In this nationally representative study of U.S. middle and high school students, we found that ONP use is highly concentrated among specific subgroups. For example, males comprised nearly 90% of frequent ONP users, which means for every one female who uses ONPs frequently, there are approximately nine males. Additionally, most current ONP users were concurrent users of other tobacco or nicotine products; only 16.6% of current users had never used another tobacco or nicotine product in 2023. While the overall prevalence of current ONP use remained low in absolute terms at 1.6% in 2023, this represents a doubling since 2021 [9].

A key finding of this study is the dual or poly-use pattern of ONP usage. In 2023, most current ONP users were also using other nicotine products. While dual use with combustible cigarettes remains high, the majority of ONP users are co-users of e-cigarettes. This strong correlation may reflect common liability, where shared risk factors like sensation seeking drive adolescents to multiple types of nicotine use [18]. Another potential explanation is that ONPs offer practical advantages over other nicotine products, including lower cost, wider availability, and nicotine satisfaction, as well as enabling complementary use. The discreet nature of ONPs, which produce no aerosol or smoke, may allow adolescents to use them in settings where smoking and vaping are prohibited. [2, 3].

Another key trend is the increase in exclusive ONP users with no lifetime use of other tobacco or nicotine products, rising from 7.4% in 2021 to 16.6% in 2023. There are several potential explanations for this phenomenon. On one hand, if ONPs are diverting youth who would otherwise have initiated use of combustible cigarettes, this trend could represent a relative harm reduction benefit [10, 12, 13, 19]. The concurrent decline in cigarette smoking to record lows lends some plausibility to this hypothesis [9]. On the other hand, if ONPs are attracting youth who would otherwise have remained away from all nicotine products, this represents an expansion of the nicotine market and a new avenue for dependence [18, 20]. Given the cross-sectional nature of this data, determining the impact of ONP will require longitudinal research to track whether these otherwise nicotine naïve users progress to other products or remain exclusive ONP users, and to quantify the balance between diversion and initiation [21].

This study has several strengths, including the use of a large, nationally representative sample and rigorous weighting procedures. However, there are some limitations. First, the cross-sectional design cannot provide evidence for causal inferences regarding the diversion or initiation effects of ONPs [16, 17]. Second, self-reported data may be subject to misclassification [22]. For example, some students may have confused ONPs with snus. A sensitivity analysis excluding snus from the “other tobacco or nicotine products” composite showed that the association with ONP use was attenuated but remained substantial which suggests that while some of the observed association may reflect misclassification, the strong residual association indicates genuine co-use of ONPs with other tobacco or nicotine products. Alternatively, the stronger association when snus is included may reflect shared preferences for oral nicotine products among certain users. Third, while the trend during 2021–2023 were analyzed, the microdata for 2024 were unavailable at the time of analysis, limiting the ability to model the most recent year’s predictors [9].

In conclusion, while the prevalence of ONP use among U.S. youth remains lower than vaping, it is increasingly observed among adolescents both with and without lifetime experience of other tobacco or nicotine products, in the context of a rapidly emerging market and limited regulation. CDC has released 2024 NYTS headline estimates, but the 2024 public-use microdata were not available at the time of analysis. In the absence of future surveillance data, our findings provide the most recent evidence on the co-use profile of current ONP users and establish a baseline for future updates as and when further data become available. Regulatory strategies should focus on reducing youth attraction to ONPs, while further research is needed to clarify their potential harm reduction or initiation role.

## Data Availability

All data produced are available online at NYTS: https://www.cdc.gov/tobacco/about-data/surveys/national-youth-tobacco-survey.html

---

**Supplementary Table 1.**
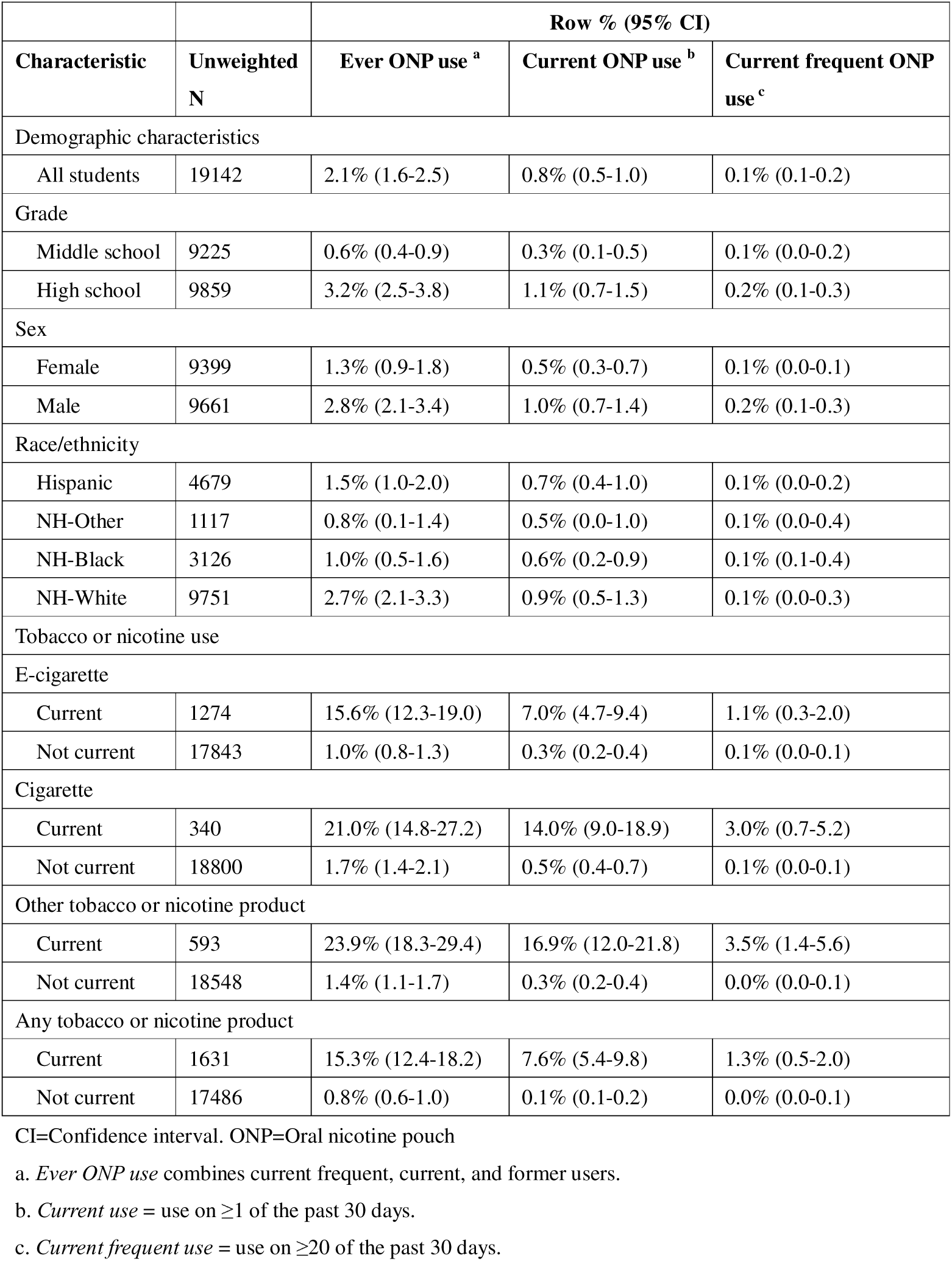
Weighted prevalence of oral nicotine pouch (ONP) use among U.S. middle and high school students, National Youth Tobacco Survey 2021.

**Supplementary Table 2.**
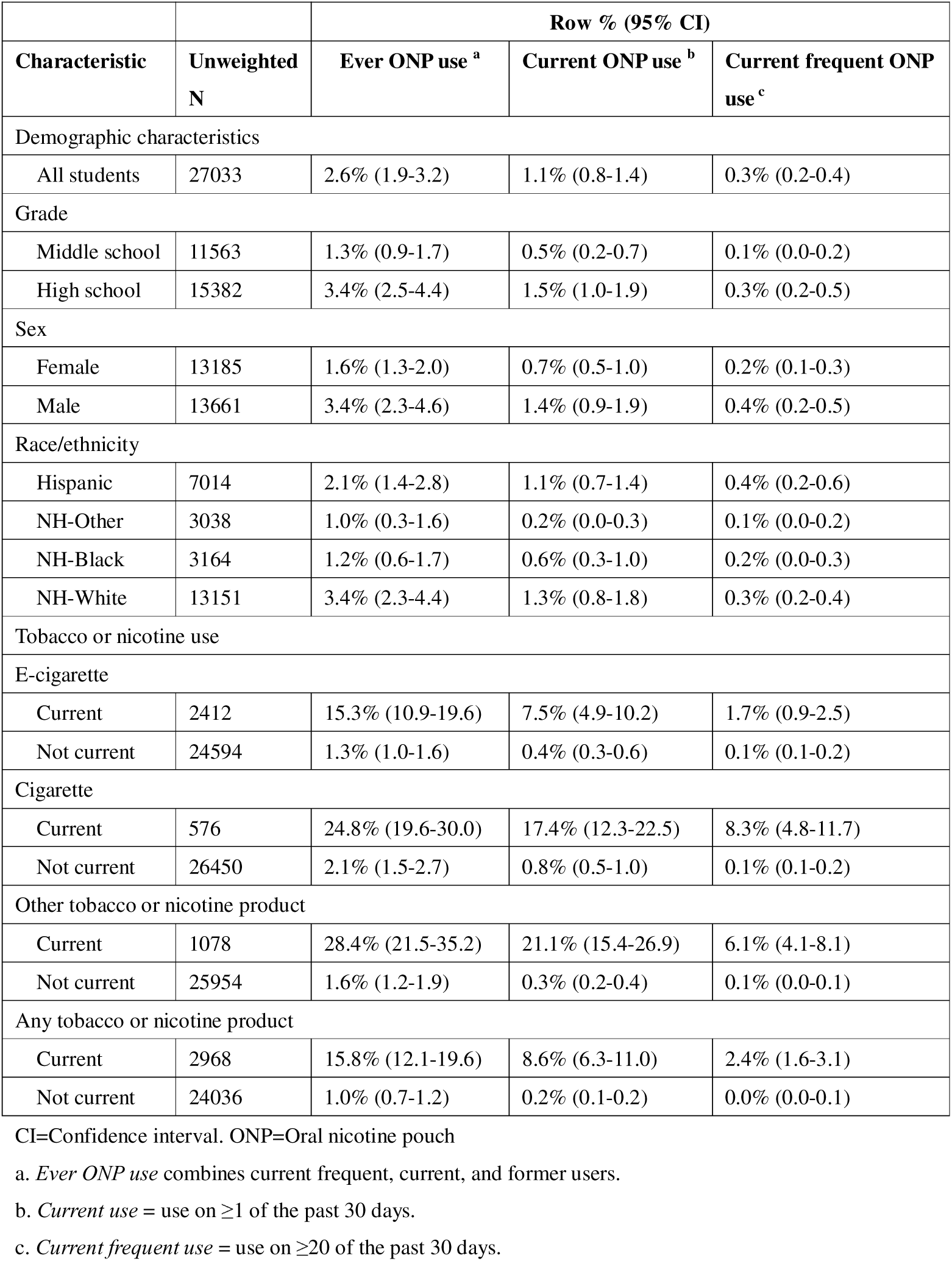
Weighted prevalence of oral nicotine pouch (ONP) use among U.S. middle and high school students, National Youth Tobacco Survey 2022.

**Supplementary Table 3.**
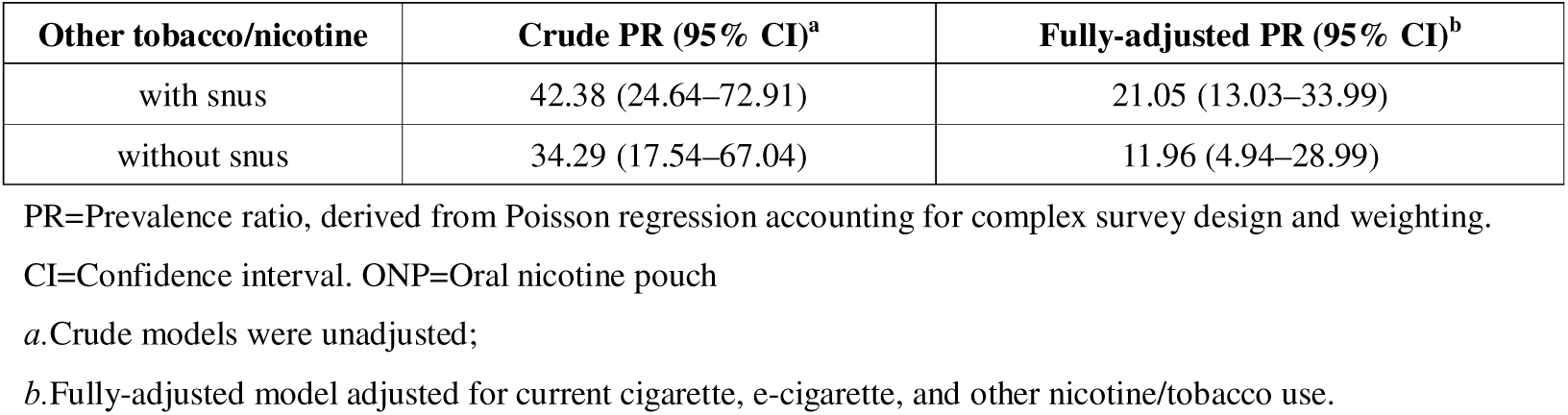
Sensitivity analysis for snus.

