## Supplementary table 1, 2, and 3 for "Oral Nicotine Pouch Use Among U.S. Middle and High School Students, 2021-2023"

**Supplementary Table 1.** Weighted prevalence of oral nicotine pouch (ONP) use among U.S. middle and high school students, National Youth Tobacco Survey 2021.

|  |  | **Row % (95% CI)** | | |
| --- | --- | --- | --- | --- |
| **Characteristic** | **Unweighted N** | **Ever ONP use ^a^** | **Current ONP use ^b^** | **Current frequent ONP use ^c^** |
| Demographic characteristics | | | | |
| All students | 19142 | 2.1% (1.6-2.5) | 0.8% (0.5-1.0) | 0.1% (0.1-0.2) |
| Grade | | | | |
| Middle school | 9225 | 0.6% (0.4-0.9) | 0.3% (0.1-0.5) | 0.1% (0.0-0.2) |
| High school | 9859 | 3.2% (2.5-3.8) | 1.1% (0.7-1.5) | 0.2% (0.1-0.3) |
| Sex | | | | |
| Female | 9399 | 1.3% (0.9-1.8) | 0.5% (0.3-0.7) | 0.1% (0.0-0.1) |
| Male | 9661 | 2.8% (2.1-3.4) | 1.0% (0.7-1.4) | 0.2% (0.1-0.3) |
| Race/ethnicity | | | | |
| Hispanic | 4679 | 1.5% (1.0-2.0) | 0.7% (0.4-1.0) | 0.1% (0.0-0.2) |
| NH-Other | 1117 | 0.8% (0.1-1.4) | 0.5% (0.0-1.0) | 0.1% (0.0-0.4) |
| NH-Black | 3126 | 1.0% (0.5-1.6) | 0.6% (0.2-0.9) | 0.1% (0.1-0.4) |
| NH-White | 9751 | 2.7% (2.1-3.3) | 0.9% (0.5-1.3) | 0.1% (0.0-0.3) |
| Tobacco or nicotine use | | | | |
| E-cigarette | | | | |
| Current | 1274 | 15.6% (12.3-19.0) | 7.0% (4.7-9.4) | 1.1% (0.3-2.0) |
| Not current | 17843 | 1.0% (0.8-1.3) | 0.3% (0.2-0.4) | 0.1% (0.0-0.1) |
| Cigarette | | | | |
| Current | 340 | 21.0% (14.8-27.2) | 14.0% (9.0-18.9) | 3.0% (0.7-5.2) |
| Not current | 18800 | 1.7% (1.4-2.1) | 0.5% (0.4-0.7) | 0.1% (0.0-0.1) |
| Other tobacco or nicotine product | | | | |
| Current | 593 | 23.9% (18.3-29.4) | 16.9% (12.0-21.8) | 3.5% (1.4-5.6) |
| Not current | 18548 | 1.4% (1.1-1.7) | 0.3% (0.2-0.4) | 0.0% (0.0-0.1) |
| Any tobacco or nicotine product | | | | |
| Current | 1631 | 15.3% (12.4-18.2) | 7.6% (5.4-9.8) | 1.3% (0.5-2.0) |
| Not current | 17486 | 0.8% (0.6-1.0) | 0.1% (0.1-0.2) | 0.0% (0.0-0.1) |

CI=Confidence interval. ONP=Oral nicotine pouch

a. *Ever ONP use* combines current frequent, current, and former users.

b. *Current use* = use on ≥1 of the past 30 days.

c. *Current frequent use* = use on ≥20 of the past 30 days.

**Supplementary Table 2.** Weighted prevalence of oral nicotine pouch (ONP) use among U.S. middle and high school students, National Youth Tobacco Survey 2022.

|  |  | **Row % (95% CI)** | | |
| --- | --- | --- | --- | --- |
| **Characteristic** | **Unweighted N** | **Ever ONP use ^a^** | **Current ONP use ^b^** | **Current frequent ONP use ^c^** |
| Demographic characteristics | | | | |
| All students | 27033 | 2.6% (1.9-3.2) | 1.1% (0.8-1.4) | 0.3% (0.2-0.4) |
| Grade | | | | |
| Middle school | 11563 | 1.3% (0.9-1.7) | 0.5% (0.2-0.7) | 0.1% (0.0-0.2) |
| High school | 15382 | 3.4% (2.5-4.4) | 1.5% (1.0-1.9) | 0.3% (0.2-0.5) |
| Sex | | | | |
| Female | 13185 | 1.6% (1.3-2.0) | 0.7% (0.5-1.0) | 0.2% (0.1-0.3) |
| Male | 13661 | 3.4% (2.3-4.6) | 1.4% (0.9-1.9) | 0.4% (0.2-0.5) |
| Race/ethnicity | | | | |
| Hispanic | 7014 | 2.1% (1.4-2.8) | 1.1% (0.7-1.4) | 0.4% (0.2-0.6) |
| NH-Other | 3038 | 1.0% (0.3-1.6) | 0.2% (0.0-0.3) | 0.1% (0.0-0.2) |
| NH-Black | 3164 | 1.2% (0.6-1.7) | 0.6% (0.3-1.0) | 0.2% (0.0-0.3) |
| NH-White | 13151 | 3.4% (2.3-4.4) | 1.3% (0.8-1.8) | 0.3% (0.2-0.4) |
| Tobacco or nicotine use | | | | |
| E-cigarette | | | | |
| Current | 2412 | 15.3% (10.9-19.6) | 7.5% (4.9-10.2) | 1.7% (0.9-2.5) |
| Not current | 24594 | 1.3% (1.0-1.6) | 0.4% (0.3-0.6) | 0.1% (0.1-0.2) |
| Cigarette | | | | |
| Current | 576 | 24.8% (19.6-30.0) | 17.4% (12.3-22.5) | 8.3% (4.8-11.7) |
| Not current | 26450 | 2.1% (1.5-2.7) | 0.8% (0.5-1.0) | 0.1% (0.1-0.2) |
| Other tobacco or nicotine product | | | | |
| Current | 1078 | 28.4% (21.5-35.2) | 21.1% (15.4-26.9) | 6.1% (4.1-8.1) |
| Not current | 25954 | 1.6% (1.2-1.9) | 0.3% (0.2-0.4) | 0.1% (0.0-0.1) |
| Any tobacco or nicotine product | | | | |
| Current | 2968 | 15.8% (12.1-19.6) | 8.6% (6.3-11.0) | 2.4% (1.6-3.1) |
| Not current | 24036 | 1.0% (0.7-1.2) | 0.2% (0.1-0.2) | 0.0% (0.0-0.1) |

CI=Confidence interval. ONP=Oral nicotine pouch

a. *Ever ONP use* combines current frequent, current, and former users.

b. *Current use* = use on ≥1 of the past 30 days.

c. *Current frequent use* = use on ≥20 of the past 30 days.

**Supplementary Table 3.** Sensitivity analysis for snus

| **Other tobacco/nicotine** | **Crude PR (95% CI)^a^** | **Fully-adjusted PR (95% CI)^b^** |
| --- | --- | --- |
| with snus | 42.38 (24.64–72.91) | 21.05 (13.03–33.99) |
| without snus | 34.29 (17.54–67.04) | 11.96 (4.94–28.99) |

PR=Prevalence ratio, derived from Poisson regression accounting for complex survey design and weighting. CI=Confidence interval. ONP=Oral nicotine pouch

*a.*Crude models were unadjusted;

*b.*Fully-adjusted model adjusted for current cigarette, e-cigarette, and other nicotine/tobacco use.
